# Viral Kinetics and Antibody Responses in Patients with COVID-19

**DOI:** 10.1101/2020.03.24.20042382

**Authors:** Wenting Tan, Yanqiu Lu, Juan Zhang, Jing Wang, Yunjie Dan, Zhaoxia Tan, Xiaoqing He, Chunfang Qian, Qiangzhong Sun, Qingli Hu, Honglan Liu, Sikuan Ye, Xiaomei Xiang, Yi Zhou, Wei Zhang, Yanzhi Guo, Xiu-Hua Wang, Weiwei He, Xing Wan, Fengming Sun, Quanfang Wei, Cong Chen, Guangqiang Pan, Jie Xia, Qing Mao, Yaokai Chen, Guohong Deng

## Abstract

**Background:** A pandemic of coronavirus disease 2019 (COVID-19) caused by severe acute respiratory syndrome coronavirus 2 (SARS-CoV-2) has been spreading over the world. However, the viral dynamics, host serologic responses, and their associations with clinical manifestations, have not been well described in prospective cohort.

**Methods:** We conducted a prospective cohort and enrolled 67 COVID-19 patients admitting between Jan 26 and Feb 5, 2020. Clinical specimens including nasopharyngeal swab, sputum, blood, urine and stool were tested periodically according to standardized case report form with final follow-up on February 27. The routes and duration of viral shedding, antibody response, and their associations with disease severity and clinical manifestations were systematically evaluated. Coronaviral particles in clinical specimens were observed by transmission electron microscopy (TEM).

**Results:** The median duration of SARS-CoV-2 RNA shedding were 12 (3-38), 19 (5-37), and 18 (7-26) days in nasopharyngeal swabs, sputum and stools, respectively. Only 13 urines (5.6%) and 12 plasmas (5.7%) were viral positive. Prolonged viral shedding was observed in severe patients than that of non-severe patients. Cough but not fever, aligned with viral shedding in clinical respiratory specimens, meanwhile the positive stool-RNA appeared to align with the proportion who concurrently had cough and sputum production, but not diarrhea. Typical coronaviral particles could be found directly in sputum by TEM. The anti-nucleocapsid-protein IgM started on day 7 and positive rate peaked on day 28, while that of IgG was on day 10 and day 49 after illness onset. IgM and IgG appear earlier, and their titers are significantly higher in severe patients than non-severe patients (*p*<0.05). The weak responders for IgG had a significantly higher viral clearance rate than that of strong responders (*p*= 0.011).

**Conclusions:** Nasopharyngeal, sputum and stools rather than blood and urine, were the major shedding routes for SARS-CoV-2, and meanwhile sputum had a prolonged viral shedding. Symptom cough seems to be aligned with viral shedding in clinical respiratory and fecal specimens. Stronger antibody response was associated with delayed viral clearance and disease severity.

**Summary boxes:** *What is already known on this topic:* As a newly appearing infectious disease, early efforts have focused on virus identification, describing the epidemiologic characteristics, clinical course, prognostics for critically illed cases and mortality. Among COVID-19 cases reported in mainland China (72 314 cases, updated through February 11, 2020), 81% are mild, 14% are severe, and 5% are critical. The estimated overall case fatality rate (CFR) is 2.3%. Some case series reported had shown that SARS-CoV-2 could shed in upper/lower respiratory specimens, stools, blood and urines of patients. However, important knowledge gaps remain, particularly regarding full kinetics of viral shedding and host serologic responses in association with clinical manifestations and host factors.

*What this study adds:* The incubation period has no change after spreading out of Wuhan, and has no sex or age differences, however, children had prolonged incubation period. Due to early recognition and intervention, COVID-19 illness of Chongqing cohort is milder than that of Wuhan patients reported. This prospective cohort study on SARS-CoV-2 infection shows clearly that the viral and serological kinetics were related in duration of infection, disease severity, and clinical manifestations of COVID-19. Our data demonstrate that nasopharyngeal, sputum and stools are major shedding routes for SARS-CoV-2, and stronger NP antibody response is associated with delayed viral clearance and disease severity.

## Introduction

In December 2019, a novel coronavirus disease (COVID-19) outbreak caused by 2019-nCoV (renamed as severe acute respiratory syndrome coronavirus 2, SARS-CoV-2) started in China^1-3^, and has been pandemic over the world^4^. As a newly appearing infectious disease, early efforts have focused on virus identification, describing the epidemiologic characteristics, clinical course, prognostics for critically illed cases, and treating the sick^5^. Among COVID-19 cases reported in mainland China (72 314 cases, updated through February 11, 2020), 81% are mild, 14% are severe, and 5% are critical. The estimated overall case fatality rate (CFR) is 2.3%. No deaths were reported among mild and severe cases, but the CFR was 49.0% among critical cases^6^. Current COVID-19 outbreak is both similar and different to the prior severe acute respiratory syndrome (SARS; 2002-2003)^7^ and Middle East respiratory syndrome (MERS; 2012-ongoing) outbreaks^8^. However, important knowledge gaps remain, particularly regarding viral shedding and host serologic responses in association with clinical manifestations. Here we longitudinally assessed 67 hospitalized SARS-CoV-2-infected patients from Chongqing city (outside of the epidemic center Wuhan city), and systematically evaluate their viral and antibody kinetics in relation to duration of infection, disease severity, and clinical manifestations.

## Materials and Methods

### Patients and Study Design

In this study we followed the strengthening the Reporting of observational studies in epidemiology (STROBE) guidelines. A total of 67 patients with COVID-19 were recruited in this study from two sections (one for severe patients, another for mild or moderate patients) of Chongqing Public Health Medical Center (CPHMC), the designated hospital for COVID-19 treatment in Chongqing central area. All patients transferred into the two sections between January 26 and February 5, 2020, were enrolled in this cohort study, with final follow-up on February 27, 2020. All of them were laboratory-confirmed as having SARS-CoV-2 infection according to WHO interim guidance. Clinical specimens including nasopharyngeal swab, sputum, blood, urine and stool were collected periodically (3-6 days interval) after admission. Specimens of nine patients (among the 67 patients) who visited and admitted at the Southwest Hospital from 21 Jan to 29 Jan, 2020, were also collected before transferring into CPHMC. The flow diagram of the study design was presented in **Supplementary Figure 1**.

Epidemiological data were obtained with standardized investigation forms, which include patient demographics, symptom history, and relevant exposures. Clinical and radiological characteristics, laboratory findings, daily information regarding symptoms, clinical course, medications, patient vital signs, complications, treatment and outcomes data were obtained with standardized case report forms from the medical records of each patient. All data were checked by a physician and two clinical assistants.

This study was approved by the ethics committee of CPHMC (document no. 2020-002-01-KY, 2020-003-01-KY) and conducted in accordance with Declaration of Helsinki principles. Written informed consent was obtained from each subject.

### Molecular Assays

Viral RNA was extracted from patient specimens with Qiamp® viral RNA mini kit (QIAGEN, Hilden, Germany). All specimens were handled under biosafety cabinet according to laboratory biosafety guideline. Quantitative real-time reverse-transcriptase polymerase chain reaction (qRT-PCR) for the Orf1ab gene was performed with qRT-PCR kit (BGI-Shenzhen, China). The specimens were considered positive if the cycle threshold (C_t_) value was ≤ 38, and negative if the results were undetermined. The C_t_ values of qRT-PCR were converted into RNA copy number of SARS-CoV-2 by a standard curve based on C_t_ values of reference plasmid DNA.

### Serological Assays

Serum specific IgM and IgG antibodies were analyzed by ELISA kits (Livzon Diagnostics Inc., Zhuhai, China), which using SARS-CoV-2 nucleocapsid protein (NP) as antigen, following the instruction manual. The OD values (450-630) were measured and titer was calculated. Three negative and two positive controls were included in each plate.

### Transmission electron microscopy

Transmission electron microscopy (TEM) was performed directly for nasopharyngeal swab, sputum and stool from patients. For negative-stain TEM, the specimen supernatant was fixed with 2% paraformaldehyde, stained with 2% phosphotungstic acid on Formvar/Carbon-coated grids. For thin-section TEM, the specimen pellets were fixed with 2% paraformaldehyde for 24h and then fixed with 1% osmium tetroxide, embedded with Eponate 12 resin. Ultrathin-sections were stained with uranyl acetate and lead citrate, separately. The negative-stained grids and ultrathin sections were observed using JEM-1400 Plus (JEOL, Japan) and HT-7700 (Hitachi, Japan) TEM, respectively.

### Data Analysis

We defined illness onset as the first day of reported symptoms consistent with COVID-19. The incubation period was defined as the time from exposure to the onset of illness^9^. We constructed epidemic curves for date of exposure to illness onset and other key dates relating to epidemic identification and disease process by R software. The key time-to-event and its difference between severe and non-severe patients were estimated by fitting a log-normal, Gamma, Weibull, or a normal distribution by one-sample Kolmogorov-Smirnov test. For continuous variables we used one-sample or paired-sample t-test. For categorical variables we used Mann-Whitney U test, Chi-square test, or Fisher’s exact test. We also performed univariable and multivariable logistic regression analysis to explore the risk factors associated with the disease severity. Statistical significance was set at p < 0.05 of 2-tailed. Statistical analyses were done using the SPSS software (v13.0) and GraphPad Prism (v5.0).

### Patient and public involvement

Patients were not involved in the study design, setting the research questions, the outcome measures, or the preparation of the manuscripts.

## Results

### Cohort Description and Epidemiological Characteristics

A total of 67 SARS-CoV-2-infected patients with 1 602 clinical specimens were enrolled in this longitudinal study (see **Supplementary Figure 1** for strategies). The demographic and epidemiological characteristics were listed in **Table 1**. The median age was 49 years (range, 10-77). About half of the patients (35, 52.2%) were men. Twenty-five of the 67 patients (37.3%) had underlying diseases. The most common symptoms at illness onset were fever (46, 68.7%) and cough (48, 71.6%).

**Table 1.** Demographic and epidemiological characteristics.

|  | All patients<br>(n=67) | Severe<br>patients (n=29) | non-Severe<br>patients (n=38) | p-value |
| --- | --- | --- | --- | --- |
| **Male sex** | 35/67 (52.2) | 18/29 (62.1) | 17/38 (44.7) | 0.159 |
| **Age, years** | 49 (10-77) | 56 (21-77) | 39 (10-74) | 0.0016 |
| **Age group, years** |  |  |  | 0.043 |
| ≤18 | 3/67 (4.5 ) | 0 | 3/38(7.9) |  |
| 19-45 | 25/67 (37.3 ) | 7/29(24.1) | 18/38(47.4) |  |
| 46-65 | 27/67 ( 40.3) | 14/29(48.3) | 13/38(34.2) |  |
| ≥ 66 | 12/67 (17.9 ) | 8/29(27.6) | 4/38(10.5) |  |
| **Smoking** | 7/67 ( 10.4) | 5/29(17.2) | 2/38(5.3) | 0.225 |
| **Drinking** | 23/67 ( 34.3) | 16/29(55.2) | 7/38(18.4) | 0.002 |
| **Case origin** |  |  |  | 0.318 |
| Imported cases | 30/67(44.8) | 15/29(51.7) | 15/38(39.5) |  |
| Secondary local cases | 37/67(55.2) | 14/29(48.3) | 23/38(60.5) |  |
| **Exposure history** |  |  |  | 0.271 |
| Recent exposure in Wuhan | 27/67(40.3) | 13/29(44.8) | 14/38(36.8) |  |
| Exposure to confirmed case | 31/67(46.3) | 10/29(34.5) | 21/38(55.3) |  |
| History of travel except Wuhan | 2/67(3.0) | 2/29(6.9) | 0/38(0.0) |  |
| Exposure to person from Wuhan | 4/67(6.0) | 2/29(6.9) | 2/38(5.3) |  |
| No recognized risks | 3/67(4.5) | 2/29(6.9) | 1/38(2.6) |  |
| **Possible exposure style** |  |  |  | 0.158 |
| Epidemic area contact | 27/67(40.3) | 13/29(44.8) | 14/38(36.8) |  |
| Secondary, household contact | 26/67(38.8) | 8/29(27.6) | 18/38(47.4) |  |
| Secondary, in restaurant | 7/67(10.4) | 3/29(10.3) | 4/38(10.5) |  |
| Secondary, public area | 2/67 (3.0) | 1/29 (3.4) | 1/38 (2.6) |  |
| Secondary, on train | 1/67 (1.5) | 0 | 1/38 (2.6) |  |
| Uncertain | 4/67 (6.0) | 4/29 (13.8) | 0 |  |
| **Generation of virus transmission** |  |  |  | 0.641 |
| P1 | 28/67 (41.8) | 14/29 (48.3) | 14/38 (36.8) |  |
| P2 | 34/67 (50.7) | 13/29 (44.8) | 21/38 (55.3) |  |
| P3 | 5/67 (7.5) | 2/29 (6.9) | 3/38 (7.9) |  |
| **Number of clusters** | 10 | -- | -- |  |
| **Number of patients from clusters** | 33/67 (49.3) | -- | -- |  |
| **Incubation period, days** | 6.0 (1-15) | 5.5 (1-11) | 7.0 (1-15) | 0.302 |
| **Underlying condition** |  |  |  |  |
| None reported | 42/67 (62.7) | 12/29 (41.4) | 30/38 (78.9) |  |
| Any reported underlying condition | 25/67 (37.3) | 17/29 (58.6) | 8/38 (21.1) | 0.002 |
| Pulmonary disease | 3/67 (4.5) | 3/29 (10.3) | 0 | 0.076 |
| Diabetes | 9/67 (13.4) | 7/29 (24.1) | 3/38 (7.9) | 0.025 |
| Hypertension | 11/67 (16.4) | 7/29 (24.1) | 4/38 (10.5) | 0.187 |
| Chronic liver disease | 7/67 (10.4) | 4/29 (13.8) | 3/38 (7.9) | 0.456 |
| Cardiac disease | 2/67 (3.0) | 1/29 (3.4) | 1/38 (2.6) | 1 |
| **Symptoms before admission** |  |  |  |  |
| Absence of symptom | 1/67 (1.5) | 0 (0.0) | 1/38 (2.6) |  |
| Any reported symptoms | 66/67 (98.5) | 29/29 (100.0) | 37/38 (97.4) | 0.379 |
| Fever | 46/67 (68.7) | 25/29 (86.2) | 21/38 (55.3) | 0.007 |
| Cough | 48/67 (71.6) | 20/29 (69.0) | 28/38 (73.7) | 0.671 |
| Short breath | 17/67 (25.4) | 11/29 (37.9) | 6/38 (15.8) | 0.039 |
| Rhinocleisis or Rhinorrhea or Sneeze | 7/67 (10.4) | 2/29 (6.9) | 5/38 (13.2) | 0.406 |
| Fatigue | 20/67 (29.9) | 7/29 (24.1) | 13/38 (34.2) | 0.372 |
| Muscle Soreness | 6/67 (9.0) | 2/29 (6.9) | 4/38 (10.5) | 0.606 |
| Laryngopharynx discomfort | 9/67 (13.4) | 3/29 (10.3) | 6/38 (15.8) | 0.517 |
| Vomit or Diarrhea | 5/67 (7.5) | 1/29 (3.4) | 4/38 (10.5) | 0.379 |
| Others | 19/67 (28.4) | 7/29 (24.1) | 12/38 (31.6) | 0.503 |

Data are median (range) or n/N (%), where N is the total number of patients with available data.

The epidemic curve of the onset of illness among cohort patients indicate a significant decrease in the number of both imported and local cases in Chongqing metropolitan area after Wuhan City shutdown since Jan 23, 2020 (**Supplementary Figure 2**). Twenty-six (38.8%) patients were infected by household contact, 33 (49.3%) were from 10 familial clusters (**Table 1**). We estimated the median incubation period was 6.0 days (range 1-15 days). There were no gender and age difference for incubation period. However, children have a prolonged incubation period (**Supplementary Figure 3**). The key time-to-event distributions were listed in **Supplementary Table 1** and plotted in **Supplementary Figure 4**.

Among the 67 patients, 29 were classified as severe pneumonia (9 were critical cases), and 38 were non-severe pneumonia (mild or moderate pneumonia), including all three children, according to the Chinese management guideline (version 6.0) for COVID-19 (http://www.nhc.gov.cn/yzygj/s7653p/202002/8334a8326dd94d329df351d7da8aefc2/files/b218cfeb1bc54639af227f922bf6b81). Compared with non-severe patients, the severe-type patients had an older age (median 56 vs. 39 years, *p* = 0.0016), higher proportion of underlying conditions (58.6% vs. 21.1%, *p* = 0.002), and higher rate of fever (86.2% vs. 55.3%, *p* = 0.007). Severe-type patients had a prolonged time from symptom onset to infection confirmation (median 5 vs. 3 days, *p* = 0.01) and longer duration of hospitalization (median 27 vs. 20 days, *p* = 0.003) than those in non-severe patients (**Supplementary Table 1** and **Supplementary Figure 4**).

### Clinical features

The clinical characteristics of the patients after admission are shown in **Supplementary Table 2**. During hospitalization, 42.1% of non-severe had normal temperature. The laboratory findings at admission were listed in **Supplementary Table 3**. Patients with severe disease had more prominent laboratory abnormalities than those with non-severe disease. All of the 67 patients received antiviral treatment (76.1% with IFN-α 1b combined with lopinavir/ritonavir), 45 (67.2%) used oxygen support, 19 (28.4%) were given empirical antibiotic treatment. For severe patients, 29 (100%) received oxygen support, 11 (37.9%) received mechanical ventilation, 15 (51.7%) were given antibiotic treatment, and 9 (31.0%) were given systematic corticosteroid treatment (**Supplementary Table 4**). At the end of this study (Feb 27, 2020), 15 (51.7%) severe patients and 31 (81.6%) non-severe patients were discharged, and no patients had died.

**Table 2.** Characteristics and duration of SARS-CoV-2 RNA shedding in clinical specimens.

| Sample type | Total patients | Total samples | Positive (%) | Viral shedding model,<br>no./total no. (%) |  |  |  | Duration time of virus from illness onset, days<br>— median (range)* |  |  |  | Still positive after<br>NS reached<br>undetectable |
| --- | --- | --- | --- | --- | --- | --- | --- | --- | --- | --- | --- | --- |
|  |  |  |  | positive in<br>continuous<br>samples | fluctuated<br>positive | single<br>positive | negative the<br>whole<br>course | total | Severe | non-Severe | *p*-value |  |
| Nasopharyngeal swab | 67 | 377 | 63/67 (94.0) | 31/67 (46.3) | 27/67 (40.3) | 5/67 (7.4) | 4/67 (6.0) | 12 (3-38) | 14 (5-38) | 11 (3-28) | 0.054 | na |
| Sputum | 61 | 221 | 58/61 (95.1) | 50/61 (82.0) | 6/61 (9.8) | 2/61 (3.3) | 3/61 (4.9) | 19 (5-37) | 23 (6-37) | 16 (5-33) | 0.068 | 28/46 (60.9) |
| Stool | 62 | 220 | 45/62 (72.6) | 19/62 (30.6) | 5/62 (8.1) | 22/62 (35.5) | 16/62 (25.8) | 18 (7-26) | 19.5 (14-26) | 18(7-25) | 0.492 | 14/46 (30.4) |
| Urine | 64 | 231 | 12/64 (18.8) | 1/64 (1.6) | 0 | 11/64 (17.2) | 52/64 (81.2) | na | na | na | na | na |
| Plasma | 63 | 211 | 9/63 (14.3) | 1/63 (1.6) | 2/63 (3.2) | 6/63 (9.5) | 54/63 (85.7) | na | na | na | na | na |
| Any sample type | 67 | 1260 | 67/67 (100.0) | na | na | na | na | 22 (3-38) | 23 (7-38) | 20 (3-33) | 0.023 | na |

\* Duration time for nasopharyngeal swab, sputum, and stool were evaluated in patients with continuous positive samples; NS: nasopharyngeal swab; na: not applicable.

**Table 3.**
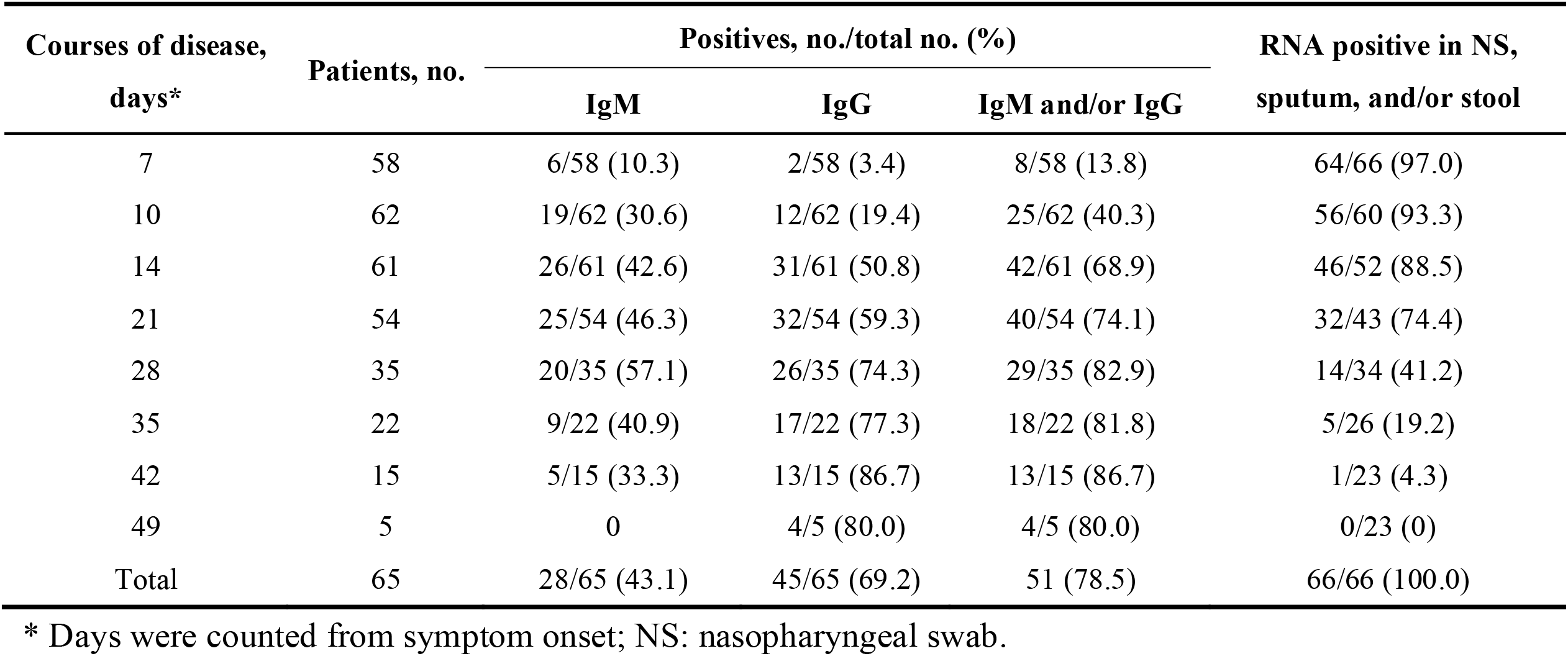
Detection of SARS-CoV-2 specific IgM and IgG antibodies in patients during the acute and early convalescent phase.

**Table 4.** SARS-CoV-2 specific IgM and IgG level and response intensity in severe and non-severe patients.

| Variable | Serum IgM level |  |  |  | Serum IgG level |  |  |  |
| --- | --- | --- | --- | --- | --- | --- | --- | --- |
|  | All patients<br>(n=58) | Severe<br>patients (n=27) | non-Severe<br>patients (n=31) | *p*-value | All patients<br>(n=54) | Severe patients<br>(n=28) | non-Severe<br>patients (n=26) | *p*-value |
| Antibodies production |  |  |  | 0.3 |  |  |  | 0.808 |
| positive | 28/58 (48.3) | 15/27 (53.6) | 13/31 (41.9) |  | 45/54 (83.3) | 23/28 (82.1) | 22/26 (84.6) |  |
| negative | 30/58 (51.7) | 12/27 (44.4) | 18/31 (58.1) |  | 9 (16.7) | 5/28 (17.9) | 4/26 (15.4) |  |
| Antibody response intensity |  |  |  | 0.017 |  |  |  | 0.032 |
| > 2 times [strong response] | 18/58 (31.0) | 13/27 (48.1) | 5/31 (16.1) |  | 12/54 (22.2) | 10/28 (35.7) | 2/26 (7.7) |  |
| 1-2 times [week response] | 10/58 (17.2) | 2/27 (7.4) | 8/31 (25.8) |  | 33/54 (61.1) | 13/28 (46.4) | 20/26 (76.9) |  |
| negative [non-response] | 30/58 (51.7) | 12/27 (44.4) | 18/31 (58.1) |  | 9/54 (16.7) | 12/27 (17.9) | 4/26 (15.4) |  |
| Antibody level in positive patients |  |  |  | 0.008 |  |  |  | 0.009 |
| > 2 times [strong response] | 18/28 (64.3) | 13/15 (86.7) | 5/13 (38.5) |  | 12/45 (26.7) | 10/23 (43.5) | 2/22 (9.1) |  |
| 1-2 times [week response] | 10/28 (35.7) | 2/15 (13.3) | 8/13 (61.5) |  | 33/45 (73.3) | 13/23 (56.5) | 20/22 (90.9) |  |
| Days of antibody first detectable from<br>illness onset in positive patients | 12.3 ± 4.4 | 11.6 ± 3.0 | 14.0 ± 5.3 | 0.156 | 14.3 ± 4.9 | 13.4 ± 4.0 | 15.3 ± 5.7 | 0.209 |
| Patients with virus clearance at day 7 after<br>antibody developed | 10/24 (41.7) | 1/13 (7.7) | 9/11 (81.8) | 0.001 | 14/34 (41.2) | 5/19 (26.3) | 9/15 (60.0) | 0.048 |

Data are mean (standard deviation) or n/N (%), where N is the total number of patients with available data.

### Dynamics of SARS-Cov-2 RNA shedding

A total of 1 260 samples from all 67 patients were collected, including 377 nasopharyngeal swab, 221 sputum, 220 stool, 231 urine and 211 plasma samples (**Table 2**). SARS-CoV-2 RNA levels in the nasopharyngeal swabs (**Fig 1A**), sputum (**Fig 1B**) and stools (**Fig 1C**) peaked in the first week, 1-20 days and 6-13 days after symptom onset, respectively, after which RNA levels typically began to decrease. Higher viral loads (inversely related to C_t_ value) were detected in the sputum than those in the nasopharyngeal and stools (peak loads about 2.3×10^9^, 2.3×10^8^ and 1.1×10^8^ copies per mililiter, respectively, **Supplementary Figure 5**). The viral loads stratified for severe and non-severe patients were also depicted (**Fig 1D, 1E**, and **1F**).

**Fig 1.**
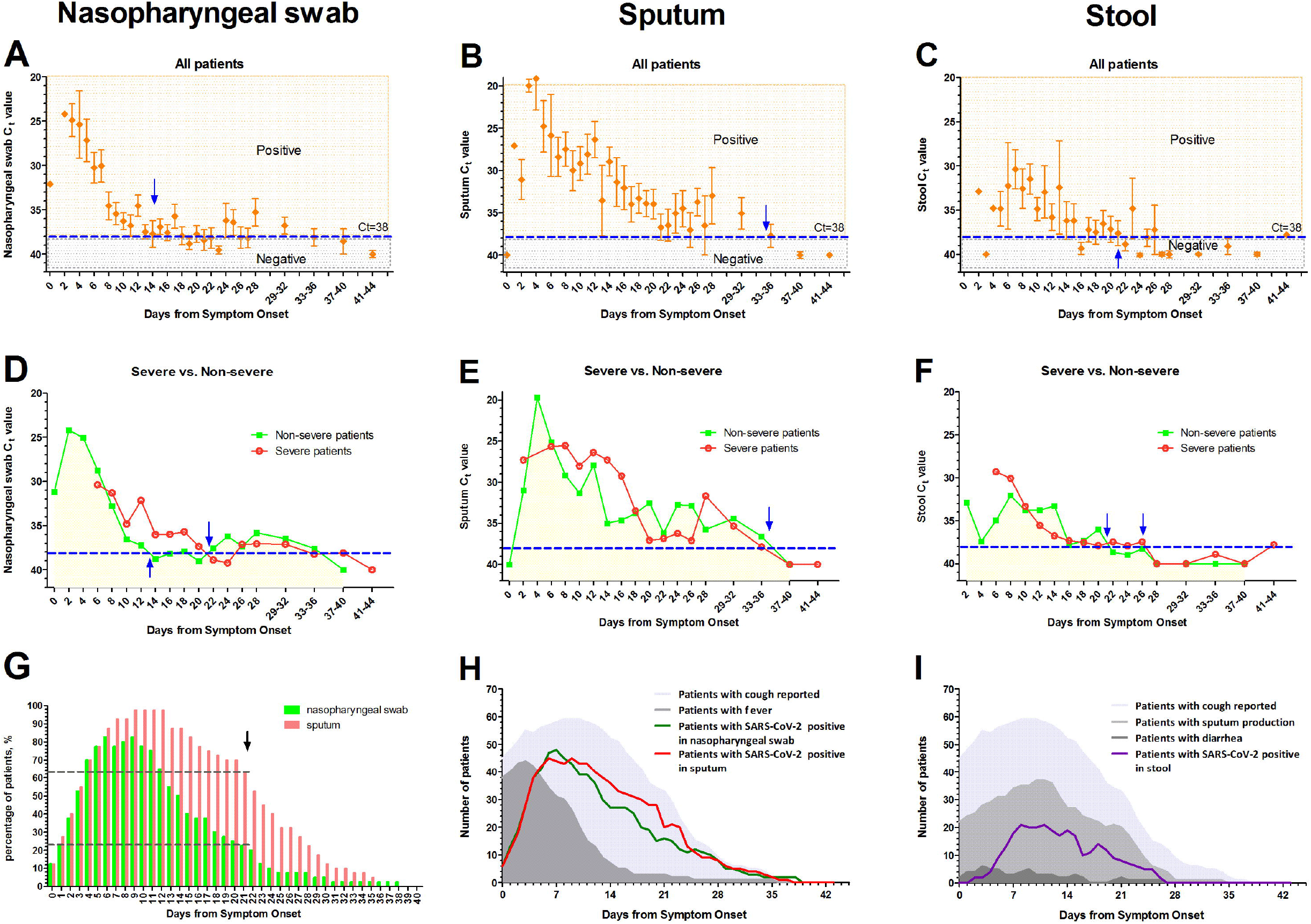
Dynamic SARS-CoV-2 Loads in Clinical Specimens and Symptom Progression over Time in a cohort with 67 Patients. The dynamic change of severe acute respiratory syndrome coronavirus 2 (SARS-CoV-2) RNA for Orf1ab gene were estimated by means of real-time reverse transcription PCR in a cohort with 67 confirmed COVID patients in nasopharyngeal swab (Panel A, with a total of 377 samples), sputum (Panel B, with a total of 221 samples), and stool (Panel C, with a total of 220 samples). The viral load was indicated by the cycle threshold (C_t_) value which was inversely related to viral RNA copy number (corresponding copy number details see in Figure S5). The blue dashed line indicates the detection limit with a C_t_ value 38. The symbol with error bar denoted the mean and its standard error of C_t_ value. Blue arrows indicated the average time to reach undetectable from symptom onset. The viral load in nasopharyngeal swab, sputum, and stool for severe and non-severe patients were also depicted in panel D, E, and F, respectively (symbol indicated the mean of C_t_ value). Panel G shows the prolonged viral shedding of sputum in 40 patients from the cohort with continuous samples both for sputum and nasopharyngeal swabs. The dark dashed lines together with the black arrows indicate an example that 25 (62.5%) patients were still RNA-positive in sputum at 21 days from illness onset, which was much higher than that in nasopharyngeal swabs (9 patients, 22.5%). Panel H and I shows the symptom (reported cough, measure fever, and diarrhea) progression and viral shedding in nasopharyngeal swabs, sputum, and stools after illness onset.

The median duration of SARS-CoV-2 RNA shedding were 12 days (range, 3-38) in nasopharyngeal swabs, 19 days (range, 5-37) in sputum and 18 days (range, 7-26) in stools (**Table 2**), and it was still detectable in any type of samples in 20.9 percent patients exceeding 30 days after symptom onset. After nasopharyngeal swabs reached undetectable among 46 patients, 28 (60.9%) and 14 (30.4%) patients were still positive for SARS-CoV-2 RNA in sputum and stools. Sputum have a longer shedding time (mean 22.0 ± 6.7 days) compared with that in nasopharyngeal swabs (mean 16.2 ± 7.2 days, *p* = 4.28×10^-7^, **Fig 1G** and **Supplementary Figure 6**). Viral shedding time was significantly longer in severe patients than non-severe patients (median 23 vs 20 days, *p* = 0.023, **Table 2** and **Supplementary Figure 7**).

Among the 231 urines and 211 plasmas collected from 67 patients, only 13 urines (5.6%) from 12 patients (18.8%) and 12 plasmas (5.7%) from 9 patients (14.3%) were positive for SARS-CoV-2 RNA (**Table 2, Supplementary Figure 8A** and **8B**). Most patients were single-point positive for urines and plasmas. There was no difference for kidney functions between urine viral positive and negative time-points (**Supplementary Figure 8C**). Additionally, six patients were confirmed as COVID-19 for SARS-CoV-2 RNA positive in sputum, bronchoalveolar lavage fluid (BALF), or stools (**Supplementary Figure 8D**).

Among patients in this COVID-19 cohort, symptom progression and SARS-CoV-2 shedding after illness onset was depicted. The numbers of patients with reported cough but not fever appeared to align with the proportion of detectable RNA both in nasopharyngeal swabs and sputum (**Fig 1H**). The numbers of patients with positive stool-RNA appeared to align with reported cough and expectoration, but not with diarrhea (**Fig 1 I**).

### Coronavirus detection by transmission electron microscopy

We found typical coronaviral particles in sputum directly by transmission electron microscopy, both for negative staining and ultrathin section preparations. As shown in **Fig 2**, typical crown-shaped coronavirus particles with spiky surface projections and an average diameter of 60-140 nm were observed.

**Fig 2.**
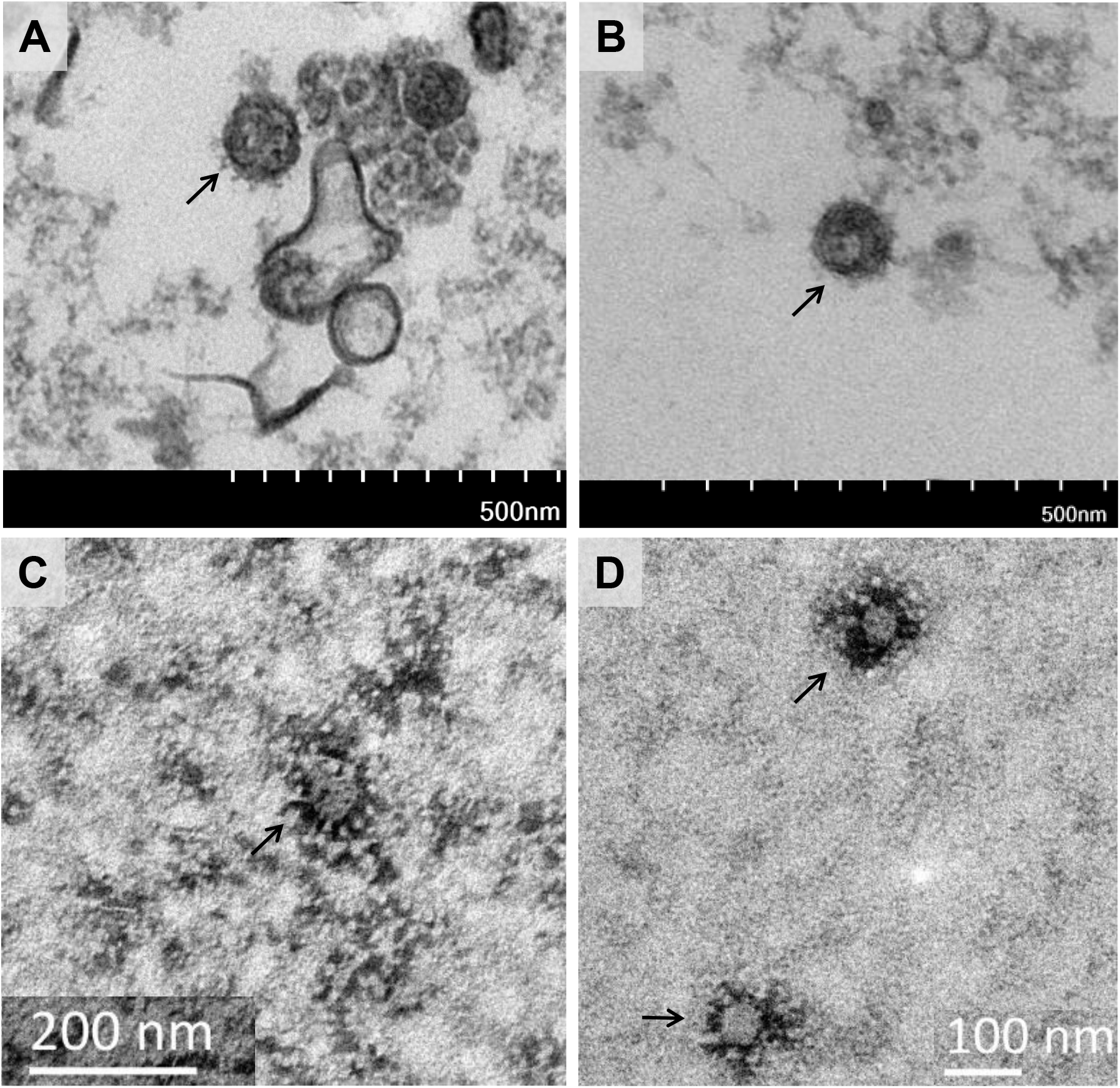
Visualization of SARS-CoV-2 with Transmission Electron Microscopy in sputum directly. Panel A and B, ultrathin-section electron-microscopy. Panel C and D, negative staining. Typical crown-shaped coronavirus particles with spiky surface projections and an average diameter of 60-140 nm in sputum from a COVID-19 patient (Patient No.14 of our cohort). The ultrathin-sections and negative-stained grids were observed by transmission electron microscopy HT-7700 (Hitachi, Japan) and JEM-1400 Plus (JEOL Corp., Japan), respectively.

### Serum antibody responses

A total of 342 sequential serum samples from 65 patients at different stages of disease progression, were tested for specific IgM and IgG antibodies to SARS-CoV-2 nucleocapsid protein. The positive rate for IgM kept increasing until 28 days (57.1%) and then decreased around 33.3 % at 42 days. The positive rate for IgG reached 74.3 % and 86.7% at 28 and 42 days, and remained (**Table 3**). The dynamic titers of serum antibodies were depicted in **Fig 3**. According to the 90 percentile of appearing time for IgM and IgG developed, we set 18 days and 21 days for IgM and IgG separately as the minimum required observation period. The patients observed less than this required time were excluded in the subsequent dynamic antibody analysis. Patients could be categorized as strong responders (peak titer > 2-fold of cutoff value), weak responders (peak titer 1-2 fold of cutoff value), and non-responders (peak titer below cutoff value). For IgM (**Fig 3A**) and IgG (**Fig 3B**), 30 (51.7%) and 9 (16.7%) were non-responders, 10 (17.2%) and 33 (61.1%) were weak responders, and 18 (31.1%) and 12 (22.2%) were strong responders (**Table 4**).

**Fig 3.**
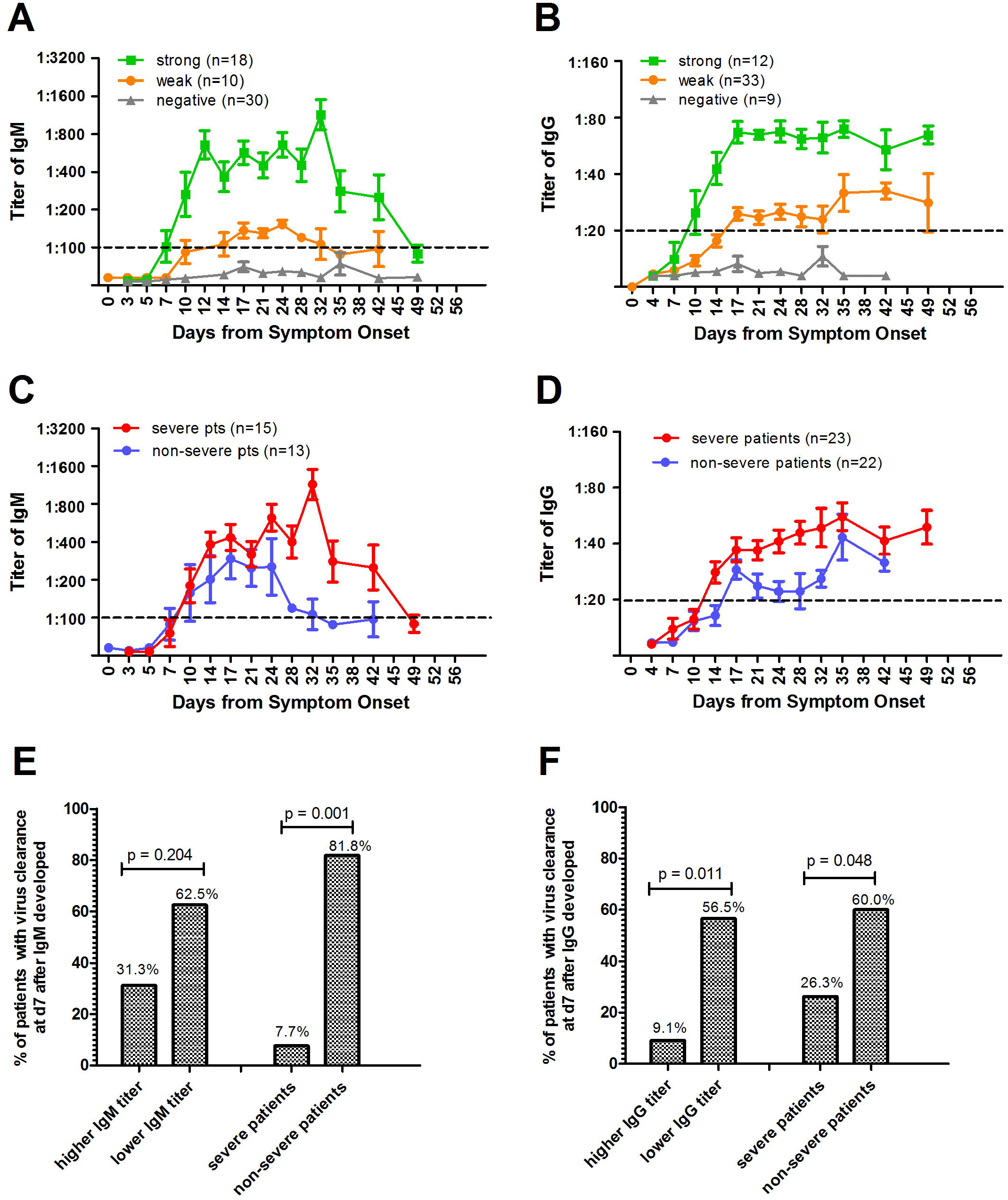
Dynamic Titers of IgM and IgG Antibodies to SARS-CoV-2 and the viral clearance in cohort. Panel A shows the specific antibody of IgM and Panel B for that of IgG. A total of 342 sequential serum samples from 65 patients at different stages were tested for antibodies and the patients observed more than 18 days for IgM (n = 58) and 21 days for IgG (n = 54) were included in the dynamic antibody analysis. The patients was set up as strong responders when peak titer > 2 fold of cutoff value, and weak responders when that of 1-2 fold. Panel C shows the different antibody response intensity in positive patients between severe and non-severe group of IgM, and Panel D for that of IgG. The dashed line denotes cutoff value for a positive result. The symbol with error bar denoted the mean and its standard error of titer. Panel E shows the comparison of viral clearance at day 7 after virus specific IgM antibody developed between severe and non-severe patients, strong and weak responders, and Panel F for that of IgG.

The proportion of strong responders is significantly higher and the proportion of weak responders is significantly lower in severe patients than that in non-severe patients, both for IgM (*p* = 0.017) and IgG (*p* = 0.032). Similarly, the titers of serum IgM and IgG were continuously significantly higher in severe patients than those in non-severe patients along with time (IgM, *p* = 0.008; IgG *p* = 0.009; **Fig 3C, Fig 3D, and Table 4**). It’s notable that the proportion for viral clearance at day 7 after antibodies appearance was significantly higher in non-severe patients than that in severe patients (for IgM, 81.8% vs. 7.7%, *p* = 0.001; for IgG, 60.0% vs. 26.3, *p* = 0.048). Furthermore, the weak responders for IgG antibodies had a significantly higher viral clearance rate (56.5%) than that (9.1%) of strong responders (p= 0.011, **Fig 3E, Fig 3F**, and **Table 4**).

### Risk factors for disease severity

In a multivariable model that included available data from 67 patients, any reported fever (OR, 17.9 [95% CI, 1.7–191.8]), underlying conditions (OR, 12.5 [95% CI, 2.0–77.6]), lesion in both lungs for radiography (OR, 6.3 [95% CI, 1.1–34.9]), bacterial infections (OR, 9.5 [95% CI, 1.3–68.0]), albumin decrease (OR, 33.7 [95% CI, 2.8–407.0]) and lactate dehydrogenase levels (OR, 31.9 [95% CI, 7.2–141.7]), and strong IgM response (OR, 9.1 [95% CI, 1.4–59.6]) were independent factors associated with the severe patients (**Table 5**).

**Table 5.** Risk factors associated with severe process in patients with COVID-19.

| Potential risk factors | Univariable |  | Multivariable |  |
| --- | --- | --- | --- | --- |
|  | OR (95%CI) | p-value | OR (95%CI) | p-value |
| **Demographics and clinical characteristics** |  |  |  |  |
| Female gender (vs male) | 0.5 (0.2 - 1.3) | 0.162 |  |  |
| Age (years) | 1.1 (1.0 - 1.1) | 0.003 |  |  |
| 19-45 | Reference |  |  |  |
| ≥18 | 0 | 0.999 | 0 | - |
| 46-65 | 2.8 (0.9 - 8.8) | 0.084 | 2.9 (0.6 - 14.8) | 0.201 |
| ≤ 66 | 5.1 (1.2 - 22.7) | 0.031 | 10.2 (0.4 - 279.4) | 0.17 |
| Drink (vs non-drink) | 5.5 (1.8 - 16.4) | 0.002 |  |  |
| Days from onset to be conformed ≥ 4d | 3.6 (1.3 - 10.2) | 0.015 | 1.8 (0.4 - 7.8) | 0.43 |
| Any reported fever (vs non-fever) | 9.8 (2.0 - 47.4) | 0.004 | 17.9 (1.7 - 191.8) | 0.017 |
| Highest temperature, °C (vs normal 37.3 °C) |  |  |  |  |
| 37.3-38.0 | 6.7 (1.2 - 36.2) | 0.028 |  |  |
| 38.1-39.0 | 15.0 (2.7 - 82.3) | 0.002 |  |  |
| >39.0 | 4.0 (0.2 - 66.86) | 0.334 |  |  |
| Underlying condition present (vs not present) | 5.3 (1.8 - 15.6) | 0.002 | 12.5 (2.0 - 77.6) | 0.007 |
| Diabetes | 5.7 (1.1 - 30.1) | 0.039 |  |  |
| Lesion in both lungs | 5.9 (1.7 - 20.5) | 0.005 | 6.3 (1.1 - 34.9) | 0.035 |
| Bacterial infection (vs not infection) | 14.6 (3.0 - 72.5) | 0.001 | 9.5 (1.3 - 68.0) | 0.024 |
| **Laboratory findings** |  |  |  |  |
| Neutrophil rate > 70% | 7.3 (2.4 - 22.0) | 4.78×10<sup>-4</sup> | 0.6 (0.1 - 7.1) | 0.676 |
| Lymphocyte rate, % (vs normal 20-40%) |  |  |  |  |
| < 20 | 7.08 (2.0 - 23.9) | 0.002 | 4.6 (0.8 - 24.6) | 0.079 |
| > 40 | 0.5 (0.1 - 2.6) | 0.401 | 0.6 (0.1 - 5.3) | 0.658 |
| Albumin < 35 g/L | 14.1 (1.7 - 120.6) | 0.016 | 33.7 (2.8 - 407.0) | 0.006 |
| Lactate dehydrogenase > 245 U/L | 20.4 (5.9 - 71.5) | 2.31×10<sup>-6</sup> | 31.9 (7.2 - 141.7) | 5.35×10<sup>-6</sup> |
| D-dimer > 0.5 mg/L | 3.8 (1.0 - 14.1) | 0.043 | 2.3 (0.2 - 27.6) | 0.521 |
| C-reactive protein > 10 mg/L | 5.4 (1.8 - 15.8) | 0.002 | 3.3 (0.4 - 24.7) | 0.245 |
| Erythrocyte sedimentation rate > 20 mm/h | 4.8 (1.5 - 15.2) | 0.008 | 3.8 (0.7 - 21.7) | 0.133 |
| **Dynamics of virus and antibody** |  |  |  |  |
| Virus clearance within 21 days from onset | 0.4 (0.1-1.3) | 0.13 |  |  |
| IgM developed within 14 days from onset | 3.7 (1.2 - 11.4) | 0.022 | 2.0 (0.2 - 18.3) | 0.523 |
| IgM level in positive patients |  |  |  |  |
| 1-2 times [week response] | Reference |  |  |  |
| > 2 times [strong response] | 10.4 (1.6 - 66.9) | 0.014 | 9.1 (1.4 - 59.6) | 0.021 |
| IgG level in positive patients |  |  |  |  |
| 1-2 times [week response] | Reference |  |  |  |
| > 2 times [strong response] | 7.7 (1.55 - 40.9) | 0.017 |  |  |

## Discussions

Our cohort from Chongqing city provides information on the epidemiology and clinical characteristics of the COVID-19 outside Wuhan, where the disease had outbreak first. We found some features of Chongqing cases were different from the early cases reported from Wuhan, China^2, 9-11^. For example, our cases were identified and admitted to hospital at earlier stage for COVID-19 than Wuhan cases, and most cases were first- or second-generation cases with clear contact history. The incubation period has no change after spreading out of Wuhan, and has no sex or age differences. However, although none of the only three children developed to severe type of disease, they had prolonged incubation period. This may have epidemiological significance and need further investigations in large-scale cohorts.

No major differences were found between the clinical characteristics of patients in this Chongqing cohort and those reported in Wuhan^2,10-12^. However, few patients had kidney injury. During one-month observation, half severe patients and most non-severe patients were discharged, which indicates milder illness of this cohort compared with relatively more severe infections of Wuhan patients reported. Through epidemic alarm from government and media, patients with fever and upper respiratory tract symptoms were asked to go to hospital at an early stage^13^.

We characterized SARS-CoV-2 viral dynamics in a hospitalized patient cohort. Our data provide important findings for this newly discovered virus infection in human. First, unlike SARS-CoV^14^ and MERS-CoV infection^15^, SARS-CoV-2 viral shedding in the nasopharyngeal swabs, sputum and stools appeared in the early phase (3-5 days from symptom onset), peaked in the first week, decreased in the second week, and persisted up to 38 days from illness onset. The viral load was highest in sputum, higher in nasopharyngeal and lower in stools. Second, SARS-CoV-2 shedding in sputum is much longer and stable than that in nasopharyngeal and stool. Third, SARS-CoV-2 RNA was just detected sparsely with low loads in the plasma and urine of minor patients. Forth, it is cough but not fever aligns with the viral shedding in nasopharyngeal and sputum. Our analysis suggests that the viral RNA shedding pattern of patients infected with SARS-CoV-2 resembles that of patients with influenza^16^ and appears different from that seen in patients infected with SARS-CoV^14^ and MERS-CoV^17^. Although viral RNA in the nasopharyngeal swab disappears quickly, testing of multiple types of samples, including nasopharyngeal, sputum and faecal samples should increase the sensitivity of the qRT-PCR assay. Interestingly, stool shedding seems to align with cough and expectoration, but not with diarrhea. Because no report demonstrates viable SARS-CoV-2 virus could be isolated from stool^18^, our data implicates the stool SARS-CoV-2 viral RNA may directly from swallowed sputum, not from infected intestinal mucosa or bile ducts. We visualized typical coronavirus in the sputum of patient directly by electron microscopy, which demonstrated the utility of this traditional technique for clinical diagnosis of SARS-CoV-2 infection.

In this study, we also determined the IgM and IgG (antibodies to SARS-CoV-2 nucleocapsid protein) dynamics in patients at acute and early convalescent phase. Although the observed profile of antibodies against SARS-CoV-2 nucleocapsid protein was consistent with common findings with regard to acute viral infectious diseases^15,19,20^, however, we have some unique findings which may be novel and important for the understanding to SARS-CoV-2 infection. First, we observed three types of antibody responses in COVID-19 patients, strong, weak and non-response. Second, we found that the earlier response, higher antibody titer and higher proportion of strong responders for IgM and IgG were significantly associated with disease severity. Third, the weak responders for IgG antibodies had a significantly higher viral clearance rate than that of strong responders. These data indicates strong antibody response is associated with disease severity, and weak antibody response is associated with viral clearance, which resembles SARS^21^ and MERS^15^. The profile of anti-SARS-CoV-2 antibodies may be helpful for the diagnosis and in epidemiologic surveys. However, the role of various antibodies relating to disease severity, immunologically directed treatment, and vaccination efficacy, deserves urgent investigation.

Nevertheless, there are some limitations for our study. First, large-scale, multi-center cohorts from other regions are needed to verify our preliminary findings. Second, the viability of virus in stools, plasma and urine, and its role in pathogenesis or transmission need to be clarified. Third, antibodies to spike and envelope proteins, and their role for protection for SARS-CoV-2 infection or reinfection are still unknown and waiting for future investigations.

## Data Availability

Relevant anonymized data will be made available on reasonable request from the corresponding author at.

## Acknowledgments

This work was partly supported by Chongqing Health Commission COVID-19 Project 2020NCPZX01, Youth Talent Medical Technology Program of PLA (17QNP010), the Chinese Key Project Specialized for Infectious Diseases (2018ZX10723203), the TMMU key project for medical research (2018XYY10), and the Southwest Hospital Medical Science Innovation Plan (SWH2018BJKJ-01, SWH2018QNLC-04). We thank for the supports the Youth Talent Program from Third Military Medical University (Tan W and Sun F) and the Academy of Medical Sciences Newton International Fellowship (Tan W). We also thank all patients that participated in this study.

## Author Contributions

WT, YL, JZ, YC and GD designed the study. CQ, QS, QH, HL, SY, JX, YC and QM screened, enrolled and clinically managed all the COVID-19 patients. WT, YL, JZ, SL, JW, ZT, XH, XX, YZ, WZ, WH, and XW collected the clinical samples and data. WT, JW, YD, XHW and YG performed the virological and serological tests. WT, FS and GD performed the data analysis. WT, YD, QW, CC, GP, YG and GD performed the electronic microscopy. WT and GD wrote the manuscript, with contributions from all the authors. WT, YC and GD get the funding for the study.

## Ethical approval

This study was approved by the ethics committee of Chongqing Public Health Medical Center (2020-002-01-KY and 2020-003-01-KY).

## Transparency statement

The lead authors affirm that the manuscript is an honest, accurate, and transparent account of the study being reported; that no important aspects of the study have been omitted; and that any discrepancies from the study as planned have been explained.

